# “PLEXIT - Therapeutic plasma exchange (TPE) for Covid-19 cytokine release storm (CRS), a retrospective propensity matched control study”

**DOI:** 10.1101/2020.07.23.20160796

**Authors:** Sultan Mehmood Kamran, Zill-e-Humayun Mirza, Arshad Naseem, Jahanzeb Liaqat, Imran Fazal, Wasim Alamgir, Farrukh Saeed, Rizwan Azam, Maryam Hussain, Muhammad Ali Yousaf, Nadeem Ashraf, Shazia Nisar, Muhammad Zafar Ali, Salman Saleem, Kaswar Sajjad, Asad Zaman, Malik Nadeem Azam, Mehmood Hussain, Raheel Iftikhar

**Affiliations:** PEMH Pakistan; PEMH; NIBMTC Pakistan

**Keywords:** COVID-19, Cytokine release storm, Therapeutic plasma exchange, Critical disease, ARDS

## Abstract

**Purpose:** To evaluate the outcome of patients with COVID-19 triggered CRS treated with Therapeutic Plasma Exchange (TPE) as compared to propensity score matched (PSM)-controls not receiving TPE.

**Material and methods:** Using PS 1:1 matching, 90 patients were assigned 2 groups (45 receiving TPE and 45 controls). Forced matching and covariate matching was done to overcome bias between two groups.

**Results:** Median age was 60 years (range 32-73 in TPE and 37-75 in non-TPE group), p= 0.325. Median duration of symptoms 7 days (range 3-22 days in TPE and 3-20 days in non-TPE), p=0.266. Disease severity in both groups was 6.6% moderate, 44.4% severe and 49% critical. Twenty-eight-day survival was significantly superior in TPE group (91.1%) as compared to PS-matched controls (61.5%), HR 0.21, 95% CI for HR 0.09-0.53, log rank 0.002. Median duration of hospitalization was significantly reduced in TPE treated group as compared to non-TPE controls 10 days and 15 days respectively (p< 0.01). CRS resolution time was also significantly reduced in TPE treated group (6 days vs. 12 days) (p< 0.001).

**Conclusion:** Use of TPE is associated with superior overall survival, early resolution of CRS and time to discharge as compared to standard therapy for COVID-19 triggered CRS.

**Highlights:**

□ Cytokine release syndrome (CRS) plays a pivotal role in pathophysiology and progression to severe and critical disease in patients with COVID-19.
□ Beyond supportive care, there are currently no proven effective treatment options (including Tocilizumab) for coronavirus disease (COVID-19)
□ Therapeutic plasma exchange (TPE) may dampen CRS of COVID-19 by removing circulating cytokines and toxins
□ TPE is the cheapest of all novel treatments available so far to treat severe to critical COVID-19
□ By intervening early with TPE, mortality and morbidity associated with COVID-19 triggered CRS can be reduced

## INTRODUCTION

Globally, more than ten million confirmed cases of COVID-19 have been reported so far, with around 0.5 million reported deaths^**1**^. Beyond supportive care, there are currently no proven effective treatment options for coronavirus disease (COVID-19)^**2**^. With around 60% mortality^**3**^ in critical cases, it is postulated that fatal outcomes of SARS-CoV-2 are associated with excessive immune response. Cytokine release syndrome (CRS) as evidenced by lymphopenia, high levels of C-reactive protein (CRP), high Ferritin, high LDH and IL-6, and significantly abnormal coagulation parameters were commonly found in severe and critically ill patients who could not survive^**3**^. Some evidence favors using steroids or IL-6 inhibitor (Tocilizumab) to dampen cytokine storm and improve survival^**4**^. As severe and critical Covid-19 patients are prone to develop sepsis, acute respiratory distress syndrome (ARDS) and/or multiple organ failure through immune dysregulation, hence, Therapeutic plasma exchange (TPE), by removing pathogenic cytokines, has also been hypothesized to have an additive role in managing early sepsis having onset less than 12 hours^**5**^. Our study was aimed to demonstrate that efficient control of the cytokine storm during early phase might be beneficial to selective patients, however, no prospective study on TPE has been conducted so far in patients with COVID-CRS.

## MATERIALS AND METHODS

### Study design and setting

It was an interventional retrospective Propensity score matched (PSM) single centre based cohort study in Pak Emirates Military Hospital Rawalpindi (PEMH), Pakistan from 1st April to 30th June 2020. This study was carried out at the Department of Pulmonology and Critical care. PEMH is the largest Covid-19 designated hospital in the country. Data of all hospitalized patients is maintained by PEMH Covid-19 Research and evaluation cell. The study was approved by Institutional Review Board.

### Definitions of study groups

Data was extracted for patients with COVID-19 admitted with or developing Cytokine release syndrome (CRS) during their admission. CRS was defined as per National guidelines for COVID-19^**6**^ given as: Fever of equal to or more than 100 F persisting > 48 hours in absence of documented bacterial infection and ANY of the following in the presence of moderate, severe or critical COVID-19 (1) Ferritin >1000 mcg/L and rising in last 24 hours (2) Ferritin >2000 mcg/L in patient requiring high flow oxygen or ventilation (3) Lymphopenia < 800 cells/ul or lymphocyte percentage <20% and two of the following (a) Ferritin >700 mcg/mL and rising in the last 24 hours (b) LDH > 300 IU (reference 140-250 IU/L) and rising in the last 24 hours (c) D-Dimer >1000ng/mL (or >1mcg/ml) and rising in the last 24 hours (d) CRP >70 mg/L (or >10 hsCRP) and rising in the last 24 hours, in absence of bacterial infection (e) If any 3 of above are present on admission, a rising trend was not required.

Severity of disease was defined as per criteria designed by WHO^**7**^. Moderate disease was defined as COVID-19 positive case with lung infiltrates < 50% of total lung fields on CXR / peripheral ground glass opacities (GGOs) on HRCT chest but no evidence of hypoxemia. Severe disease was operationally defined as COVID-19 pneumonia with evidence of hypoxemia (RR > 30/minute or PaO2 on ABGs < 80mmHg or PF ratio < 300 or lung infiltrates > 50% of the lung field). Critical disease was defined if there was COVID-19 pneumonia with evidence of either respiratory failure (PaO2 ≤ 60mmHg) or multiorgan dysfunction syndrome (MODS) measured by SOFA score > 10 or septic shock (Systolic BP less than 90 or less than 40mm Hg of baseline in hypertensive or Urine output < 0.5 ml/kg/hour).

### Inclusion and exclusion criteria

#### Inclusion criteria included

(1) COVID-19 diagnosed by Polymerase Chain Reaction (PCR) positivity for SARS-CoV2 (2) CRS at presentation or developing during hospitalization (3) 10-80 years age and both genders (4) hospital admission (5) At least 1 completed session of plasma-exchange in patients included in TPE arm (6) No other novel therapy administered.

#### Exclusion criteria were

(1) Death within 48 hours of admission (2) severe septic shock at time of admission (3) Congestive cardiac failure (EF<20%) (4) Those receiving immunotherapy, Anti-thymocyte globulin or hematopoietic stem cell transplant in recent past (5) Patients of hematological or solid organ malignancies (6) patients receiving other investigational drugs including Tocilizumab, Convalescent plasma, Remdesivir, or Mesenchymal stem cells.

### Definitions of interventions

As per Institutional COVID-19 Management Guidelines all patients of moderate, severe and critical COVID-19 received standard protocol of aspirin, anticoagulation, ulcer prophylaxis, awake Proning (if PaO2 < 80mmHg) and corticosteroids. All patients of CRS received Methylprednisolone 1 mg/kg irrespective of disease severity. In addition to standard care TPE was offered as a trial investigational therapy to willing patients with CRS. All patients were explained the investigational role of Therapeutic plasma exchange (TPE) in treatment of COVID-19. Written consent was taken from those who agreed for this treatment. TPE was performed once daily using COBE Spectra Apheresis machine (Manufacturer TERUMO BCT, INC) having continuous flow centrifugation. Venous access was achieved using an ultrasound guided double lumen catheter (Arrow – 12 FR) via femoral vein. Patient’s total blood volume was calculated as per Nadler’s formula. Anticoagulant acid dextrose ratio was 1:10 and flow rate 30-40 ml/minutes (Adjusted as per hemodynamic status). Patients’ blood pressure, pulse, oxygen saturation was monitored throughout procedure. Duration of procedure varied from 2-4 hours and 1-1.5 times total plasma volume was removed during each procedure. Replacement fluid was fresh frozen plasma (FFP) and normal saline in 2:1 respectively. All procedures were performed in intensive care or high dependency unit by Apheresis Department of PEMH. TPE was continued till recovery.

Recovery was defined by de-escalation of patients condition from severe to moderate, or from moderate to mild, plus at least 2 of the following; serum Ferritin < 1000 ug/ml (and decreasing trend on two consecutive days), serum LDH normalization, C-reactive protein > 50% fold reduction (and decreasing trend in on two consecutive days),), ALC > 1000 and PT/APTT normalization. TPE related complications were also documented.

### Study end points

Primary end point was survival outcome. Outcome measures were death or discharge. Patient discharge criteria included normalization of CRP, LDH, d-dimers and fall in serum Ferritin to <500 mcg/ml plus an afebrile period of at least 72 hours, and maintaining oxygen saturation >93% for at least 72 hours without supplemental oxygen support.

Secondary outcome measures were (1) Duration of hospitalization (2) Timing of PCR negativity (3) Time to resolution of CRS symptoms (4) Complications of TPE

### Statistical analysis

Retrospective observational studies involving therapeutic interventions are often confounded by either measured or unmeasured baseline characteristics. As a result, baseline characteristics of treated subjects differ from untreated ones. In order to account for these systematic differences, we conducted propensity score matched (PSM) analysis of patients of COVId-19 associated CRS treated with or without TPE. PS-matching was performed on cohort of patients meeting above inclusion and exclusion criteria.

For estimation of propensity score we used a logistic regression model (data matching Greedy) on NCCS statistical software v20.0.2. Number of controls was matched with TPE treatment group in 1:1 matching. Distance calculation method used was Mahalanobis distance including the propensity score, order for matching was random 1:1 and Caliper radius was set at 1*Sigma.

For ensuring comparable groups, forced matching was done for disease severity, standard care and advanced treatment at disease escalation. Co-variate matching was done for age, duration of illness, symptoms at presentation, co-morbidities, serum Ferritin, lactate dehydrogenase, d-dimers, C-reactive protein levels, absolute lymphocyte count (ALC), platelet count and oxygen requirement at time of CRS diagnosis.

Median and range was used for continuous variables while frequency and percentage were used to express categorical statistics. The Chi-square test was used to evaluate differences in categorical variables while Students t-test or Mann-Whitney U test was used to evaluate continuous variables. Kaplan-Meier test was used for survival analysis and log rank was used to compare difference in two groups. Cox-proportional hazards were used to generate hazard ratio (HRs) and 95% confidence interval (CIs) for outcome. P-value <0.05 was considered significant.

## RESULTS

### Patient Selection

Patient selection procedure is shown in **figure-1**. On initial screening of data for COVID-19 patients, 315 cases of CRS were found. After applying exclusion criteria, 280 eligible patients were included in PSM analysis. Using 1:1 matching, 45 pairs of patients were formed, treated with or without TPE.

**Figure 1:**
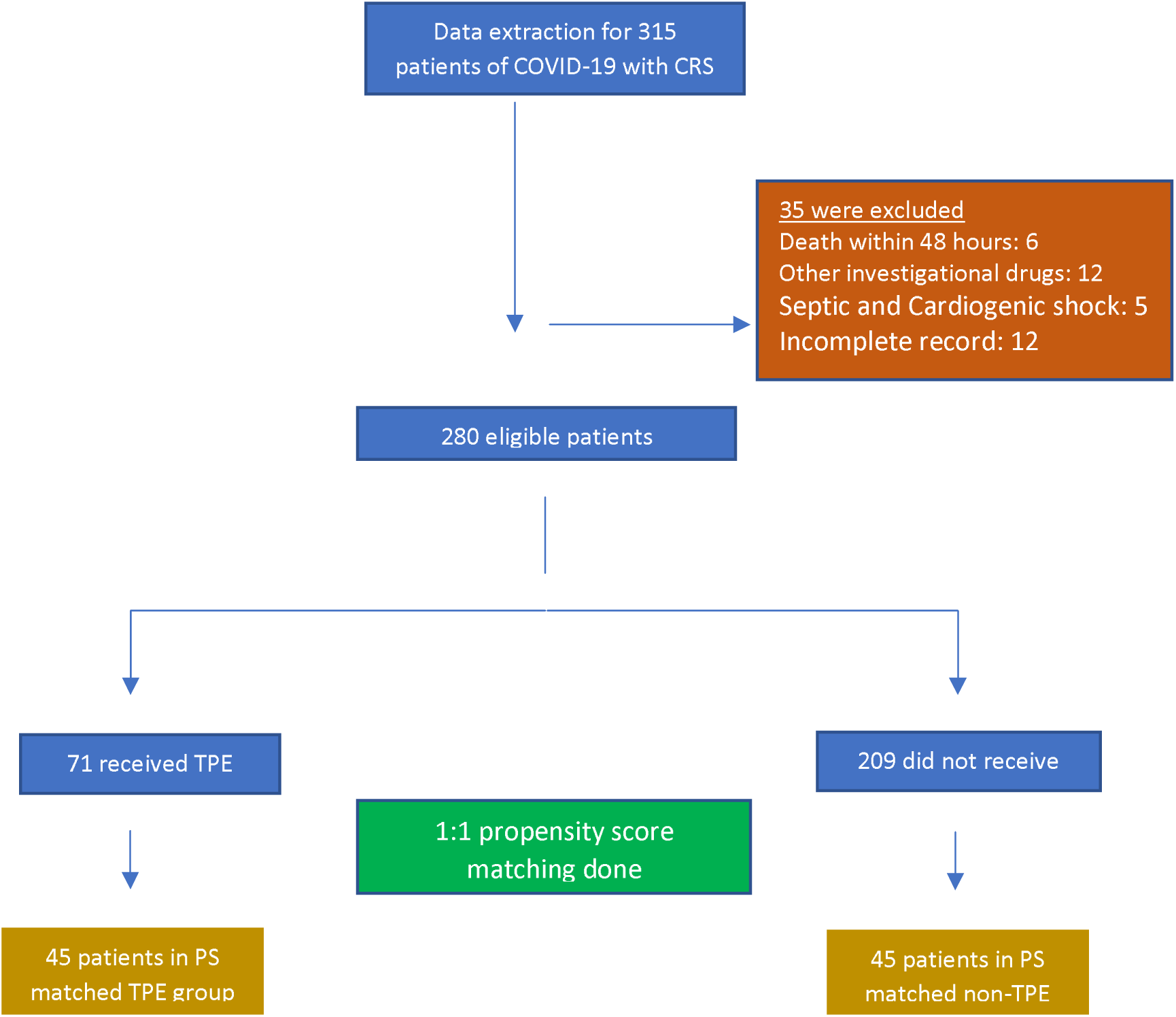
Patient selection procedure. Patient selection and matching flow chart. COVID-19, coronavirus disease 2019; TPE, therapeutic plasma exchange; PS, propensity score

### Baseline and clinical characteristics

Baseline characteristics of patients before and after matching are shown in **table 1**. Before matching there were significant differences in TPE treated and untreated groups, which were addressed after matching. The overall eligible cohort was 245/280 (87.5%) males and 35/280 (12.5%) females, with a median age of 62 years (range 20-80 years) p-value 0.05. Majority 150/280 (53.57%) had age range between 50-70 years.

**Table 1:** demographic and clinical characteristics of study group

|  | **Before PSM** |  |  | **After PSM** |  |  |
| --- | --- | --- | --- | --- | --- | --- |
| **Characteristics** | **TPE**<br>**(n= 71)** | **No TPE**<br>**(n=209)** | **P**<br>**value** | **TPE**<br>**(n= 45)** | **No TPE**<br>**(n= 45)** | **P value** |
| Age years, median<br>(range) | 60 (32-80) | 64 (20-91) | 0.05 | 60(32-73) | 60(37-75) | 0.325 |
| **Age groups, n (%)** |  |  | <0.001 |  |  | 0.690 |
| 10-30 years | 0 | 4 (1.9%) |  | - | - |  |
| 30-50 years | 19 (26.7%) | 42 (20%) |  | 8 (17.8%) | 7 (15.6%) |  |
| 50-70 years | 45 (63.3%) | 105 (50.2%) |  | 32(71.1%) | 32 (71.1%) |  |
| >70 years | 7 (10%) | 58 (27.7%) |  | 5 (11.1%) | 6 (13.3%) |  |
| **Gender, n (%)** |  |  | 0.235 |  |  | 0.306 |
| Male | 65 (92%) | 180 (86.1%) |  | 44 (97.7%) | 42 (93.3%) |  |
| Female | 6 (8%) | 29 (13.9%) |  | 1 (2.3%) | 3 (6.7%) |  |
| **Co morbidities, n(%)** |  |  | 0.137 |  |  | 0.98 |
| Obstructive air way<br>disease | 3 (4.2%) | 10 (5.5%) |  | 2(4.4%) | 2(4.4%) |  |
| IHD | 5 (7%) | 11 (5.2%) |  | 3(6.7%) | 3(6.6%) |  |
| Down syndrome | 1 (1.4%) | - |  | - | - |  |
| DM | 5 (7%) | 23 (11%) |  | 6(13.3%) | 6(13.3%) |  |
| DM+HTN | 12 (17%) | 19 (9%) |  | 2(4.4%) | 2(4.4%) |  |
| IHD+DM+HTN | 6 (8.6%) | 17 (8.1%) |  | 3(6.7%) | 3(6.7%) |  |
| HTN | 7 (9.8%) | 24 (11.4%) |  | 4 (8.88%) | 4 (8.88%) |  |
| >3 co morbidities | 1 (1.4%) | 1 (0.47%) |  | 4 (9%) | 4 (9%) |  |
| RA+ILD | - | 1 (0.47%) |  | - | - |  |
| CVA | 1 (1.4%) | 1 (0.47%) |  | - | - |  |
| CKD | 1 (1.4%) | 1 (0.47%) |  | - | - |  |
| No | 29 (40.8%) | 100 (47.8%) |  | 21 (46.7%) | 21 (46.7%) |  |
| **Clinical features n(%)** |  |  |  |  |  |  |
| Cough | 59 (83%) | 168 (80%) | 0.61 | 36(80%) | 37(82.2%) | 0.787 |
| SOB | 58(81%) | 144 (68.8%) | 0.04 | 36 (80%) | 40 (88.9%) | 0.244 |
| Duration of symptoms at<br>admission, median<br>(range) | 9 (1-25) | 5 (1-20) | <0.001 | 7(3-22) | 7 (3-20) | 0.266 |
| **Laboratory values** |  |  |  |  |  |  |
| Absolute Lymphocyte<br>count x 10<sup>9</sup>/l, median<br>(range) | 754 (200-2100) | 800<br>(230-1960) | <0.001 | 700(200-2100) | 790(230-1400) | 0.692 |
| Platelet count x 10<sup>9</sup>/l,<br>median (range) | 190(70-1100) | 205 (56-450) | 0.448 | 180(70-1100) | 187(56-450) | 0.603 |
| CRP (ug/ml), median<br>(range) | 120 (6-303) | 112 (56-390) | 0.268 | 145 (21-278) | 147 (56-260) | 0.284 |
| IL-6 , median (range) | 95 (6-400) | 68 (7-679) | 0.206 | 78 (6-400) | 104(7-178) | 0.116 |
| D-dimers, median | 400(150- | 341 (200- | 0.571 | 350(150- | 647(300- | 0.642 |
| (range) | 1700) | 1100) |  | 1700) | 1100) |  |
| Ferritin (ng/ml), median (range) | 1848(378-16942) | 1505 (71-2832) | <0.001 | 1500(336-7877) | 1410(395-4500) | 0.218 |
| ALT (IU/l), median (range) | 99(23-345) | 68(21-288) | 0.057 | 64 (31-355) | 75(28-288) | 0.744 |
| LDH (U/l), median (range) | 557 (235-1590) | 548 (273-1123) | <0.001 | 549(235-1590) | 545(273-1448) | 0.817 |
| **Cardiac biomarkers,n(%)** |  |  | 0.73 |  |  | 0.188 |
| Normal | 37 (52.1%) | 103 (49.2%) |  | 23 (51%) | 28 (62.2%) |  |
| Raised | 34 (47.9%) | 106 (50.8%) |  | 22(49%) | 17 (37.8%) |  |
| **HRCT Chest findings,n(%)** |  |  | 0.09 |  |  | 0.37 |
| Typical | 65(91.5%) | 164 (78.5%) |  | 41 (91%) | 38 (84.4%) |  |
| Indeterminate | 2(2%) | 18 (8.6%) |  | 1 (2.3%) | 1 (2.2%) |  |
| Atypical | 3(4.2%) | 23 (11%) |  | 2 (4.4%) | 6 (13.4%) |  |
| Normal | 1(1.4%) | 4 (1.9%) |  | 1 (2.3%) | - |  |
| **Lung involvement (%),n(%)** |  |  | 0.07 |  |  | 0.607 |
| <50 | 17 (23.9%) | 84 (40.2%) |  | 13 (28.8%) | 13 (28.8%) |  |
| >50 | 53 (74.7%) | 121 (57.9%) |  | 31 (69%) | 32 (71.2%) |  |
| Normal | 1(1.4%) | 4(1.9%) |  | 1 (2.2%) |  |  |
| **Oxygen support Liter/min, (range)** | 10 (2-15) | 6 (0-15) | <0.001 | 10(4-15) | 10.5(0-12) | 0.730 |
| NIV | 34 (47.8) | 59 (28.2) | <0.001 | 19 (42.2%) | 19 (42.2%) | 0.66 |
| IV | 7 (10) | 6 (2.8) | 0.01 | 3 (6.6%) | 3 (6.6%) | 1 |
| **Disease severity,n(%)** |  |  | <0.001 |  |  | 1 |
| Moderate | 4(5.6%) | 71(34%) |  | 3 (6.6%) | 3 (6.6%) |  |
| Severe | 25(35.2%) | 110(52.6%) |  | 20 (44.4%) | 20 (44.4%) |  |
| Critical | 42(59.2%) | 28(13.4%) |  | 22 (49%) | 22 (49%) |  |
| **Others treatment,n(%)** |  |  |  |  |  |  |
| Steroids | 71(100%) | 209(100%) | - | 45(100%) | 45(100%) | 1 |
| Anticoagulation | 63 (88.7%) | 178 (85.1%) | 0.453 | 45(100%) | 45(100%) | 1 |
| HCQ | 7(10%) | 24 (11.4%) | 0.76 | 2 (4.4%) | 2 (4.4%) | 1 |
| Positive Day 7 PCR (67 and 189 evaluable) n(%) | 28 (41.7%) | 80 (42.2%) | 0.93 | 14(31%) | 15(33.3%) | 0.82 |
| Positive Day 14 PCR (32 and 152 evaluable),n(%) | 4 (12.5%) | 25 (19.6%) | 0.57 | 3(6.6) | 3(6.60 | 1 |
| Discharge days, median (range) | 10 (3-24) | 13(5-30) | 0.034 | 10(4-37) | 15(7-45) | 0.01 |
| Time for CRS resolution,median(range) | 7(3-20) | 11 (5-30) | 0.04 | 6(2-23) | 12(5-42) | 0.001 |

Regarding chronic health conditions, 129/280 (46.07%) had no co morbids. Hypertension (HTN) and Diabetes Mellitus (DM) either alone or in combination with other illnesses were the first and second most common co morbids respectively (HTN; 85/280 (30.35%) and DM; 82/280 (29.28%). HRCT chest showed typical COVID appearance in 229/280 (81.78%) with 174/280 (62.14%) having > 50% lung involvement.

Total of 71 patients received TPE as compared to 209 patients without TPE. Before PSM analysis, two groups differed significantly with respect to different demographic and clinical features as in **table 1**. After PSM, two groups (TPE versus non TPE) of 45 patients each had comparable characteristics.

Regarding PS-matched cohort (n=90), 71.1% (32 TPE + 32 non-TPE) had age range between 50-70 years (p-value 0.690) with 46.7% (21 TPE + 21 non-TPE), (p-value 0.98) having no co morbidities. HRCT chest had typical appearance for COVID-19 in 79/90 (87.7%) cases and >50% lung involvement in 63/90 (70%). Among 44/90 (48.8%) ventilated patients, 38/90 (42.2%) patients received Continuous positive airway pressure (CPAP) and 6/90 (6.6%) patients required mechanical ventilation.

The Cohort treated with TPE received a median of 2.25 sessions (range 1-5). Only two patients developed TPE related complications (Femoral artery puncture, Thrombophlebitis of femoral vein with DVT), which were managed optimally. Overall survival was significantly superior in TPE group (91.1%) 95% CI 38.33-44.76) as compared to PS-matched controls (61.5%) HR 0.21, 95% CI for HR 0.09-0.53, log rank 0.002. (p<0.001) **(figure-2)**.

**Figure 2:**
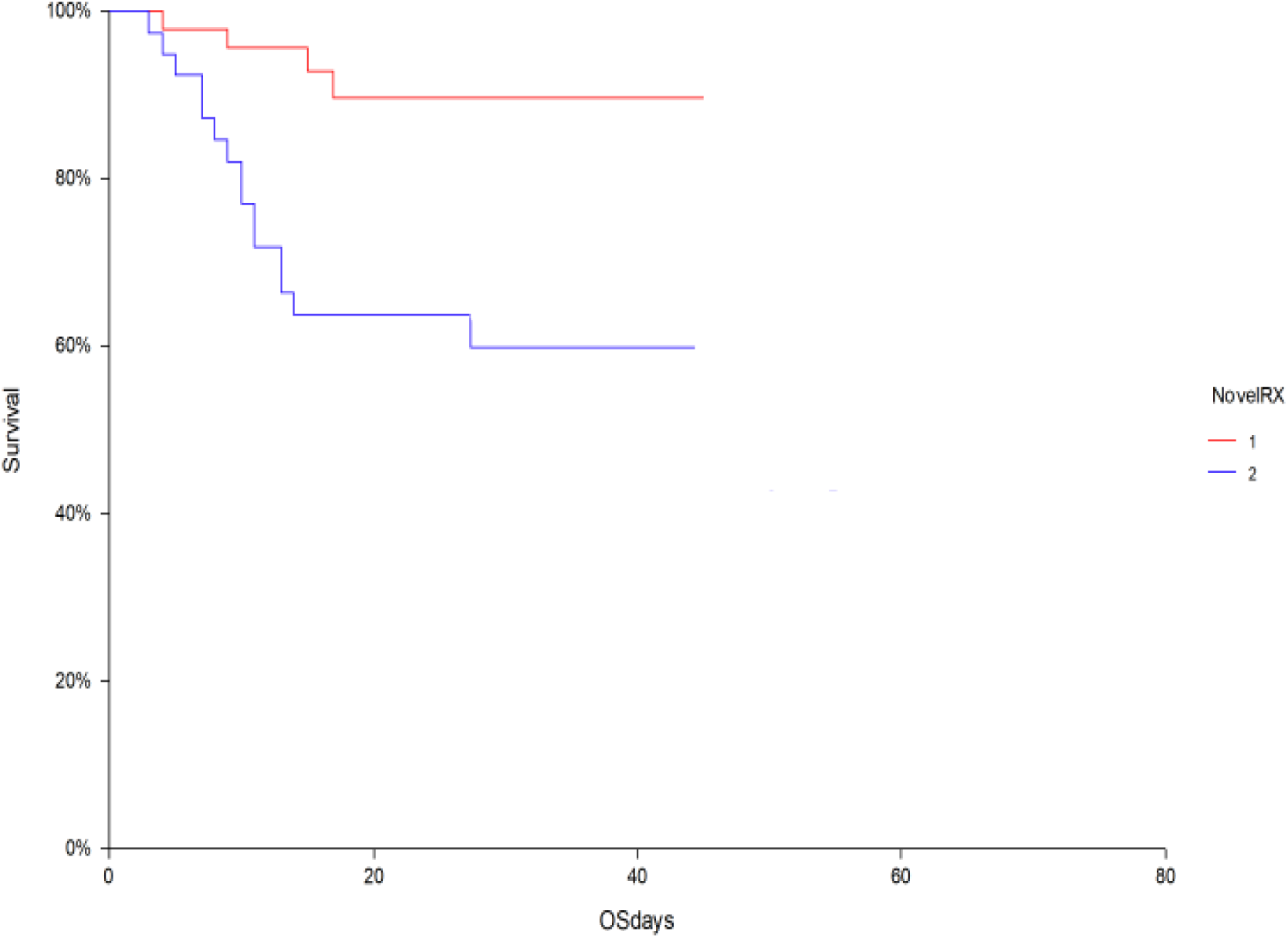
Survival Analysis. Overall survival in TPE (91.1%) 95% CI 38.33-44.76 versus PSM control (61.5%) 95% CI 29.9-48.43, log rank 0.002.

Cox regression analysis was performed to analyze effect of covariate on survival outcome in both groups. After adjusting for age, co-morbids, disease severity and duration of symptoms, Overall survival (OS) in TPE group remained superior to PSM control group (p<0.001), HR 0.21 and 95% CI 0.07-.636.

Regarding effect of co-morbids; in TPE group (n=45), Overall survival (OS) was 100% for 21 patients without co-morbidities as compared to 83.4% for 24 patients with co-morbidities. In patients not receiving TPE (n=45), OS was 77% for 21 patients without co-morbidities and 46% in 24 patients with co-morbids (p value=0.023).

Out of 45 patients in each studied group, 3 patients had moderate, 20 had severe and 22 had critical Covid-19 in each cohort respectively. Overall survival for patients with moderate, severe and critical COVID-19 was 100%, 100% and 81.8% for TPE group as compared to 100%, 90% and 40.9% for patients not receiving TPE (log rank 0.002) **(figure-3)**.

**Figure 3:**
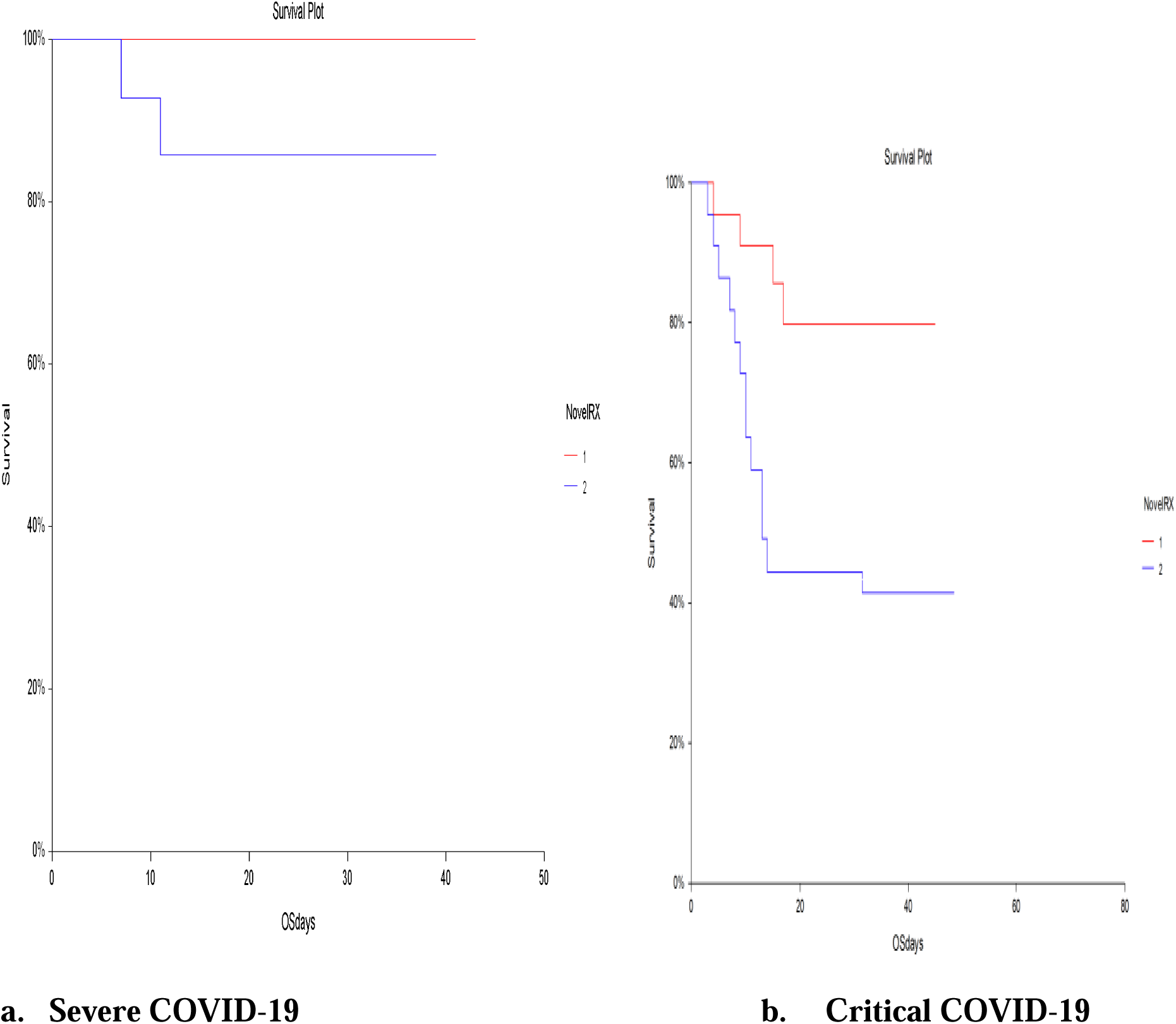
Survival comparison in severe and critical disease. Survival comparison in severe and critical Covid-19 between TPE and non-TPE cohorts (a) For severe COVID-19 patients, OS of 100% and 90% (p=0.08) in TPE and non TPE patients respectively (b) For critical COVID-19 patients, OS of 81.8% and 40.9% (p=0.007) in TPE and non TPE patients respectively

Time of resolution of CRS was significantly reduced in TPE group. From time of admission, cumulative incidence for normalization of CRS at day 15 was 90% in TPE group vs. 50% for PS-matched controls. Gray’s test was applied to cater for completing risk, difference was statistically significant (p < 0.001) **(Figure-4)**.

**Figure 4:**
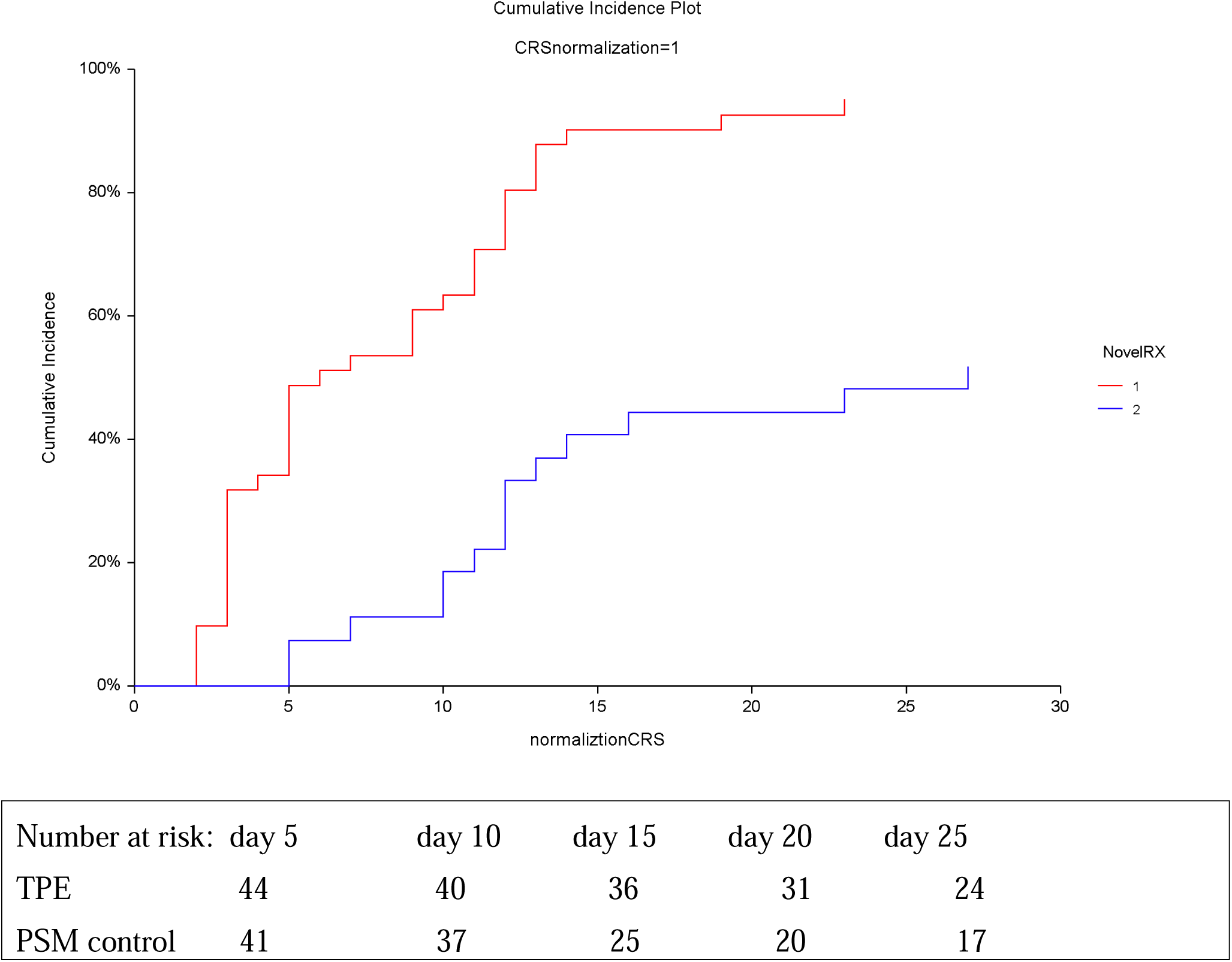
Time to CRS resolution. Cumulative incidence of normalization of CRS in TPE versus PS-matched controls.

Overall, 108/280 (38.6%) patients remained PCR positive on day 7 and 29/280 (10.35%) on day 14 with or without TPE. Median duration of hospitalization was significantly reduced in TPE treated group as compared to non-TPE controls (10 days vs. 15 days (p< 0.01).

## DISCUSSION

This study demonstrates that addition of TPE (as a single novel therapy) to the standard treatment (inclusive of steroids) for moderate, severe and critical COVID-19 with CRS is associated with significant survival benefit especially in critical disease and it remarkably decreases duration of hospitalization and resolution of CRS. However, PCR positivity at day 7 and 14 remained unchanged with addition of TPE. This is to be expected since evidence suggests presence of non-replicable viral nucleic acid material only, after day 10 of onset of illness, being picked up by the PCR**^8, 9^**.

To neutralize more comprehensively for biases associated with the selection of a particular mode of treatment, we stringently matched TPE and standard treatment care in both groups using a PSM analysis. The strength of conclusion stems from the fact that 15 variables in both arms had to be matched before analysis.

TPE appears promising as an investigational therapy for several convincing reasons. First, TPE^**10**^ has been used in secondary HLH (III,2C), thrombotic microangiopathy secondary to various causes (various categories and strengths of evidence) and septic shock (III,2B) and any/all of these pathologies may be present in severe to critical COVID-19. For instance, not only Cytokine profile of severe Covid-19 closely resemble secondary hemophagocytic lymphohistiocytosis (sHLH)^**11**^ but also it is associated with venous and arterial thromboembolic complications^**12**^ and septic shock^**13**^, Therefore, it was hypothesized that TPE will be similarly beneficial if used in COVID-19 triggered CRS. Second, TPE has been used previously for the management of severe infections as well, such as 2009 HIN1 influenza A^**14**^, sepsis and multiorgan failure with a trend towards improved survival (Knaup et al., 2018^**5**^), (Busund et al.,2002^**15)**^ and (Rimmer et al., 2014^**16)**^. Third, TPE has been proposed as a possible supportive treatment for fulminant COVID-19 infection by Keith et al., 2020^**17**^ and has been shown to be effective in a few case reports of Covid-19 (Shi et al., 2020^**18**^; Zhang et al., 2020^**19**^). However, as per Ma et al, it’s benefit in Covid-19 should be expected only in macrophage activation syndrome, or sepsis complicated with MODS (Ma et al., 2020)^**20**^.

As compared to other novel therapies, TPE facility is the easiest and cheapest to institute in a resource constrained country in global pandemic times. A key factor in its success was to start TPE early when pro-inflammatory cytokines were high (Yang et al., 2020)^**3**^. In COVID-19 the inflammatory cytokines were significantly higher around 2^nd^ week of illness (Wan et al. 2020)^**21**^. Thus, early recognition of CRS, with early initiation of TPE resulted in better clinical outcomes.

The first reported study to our knowledge on use of TPE in Covid-19 was carried out retrospectively on invasively ventilated patients receiving > 2 vasopressors by Keith et al^**17**^ which showed the greatest mortality benefit with TPE in these patients, (47.8% mortality vs. 81.3% mortality, p = 0.05). In comparison, our study having larger sample size and done at various stages of illness (moderate, severe and critical cases), with only 6 patients on mechanical ventilation included, showed significant benefit of TPE in severe and critical disease at a stage short of invasive ventilation. Out of 6 patients on mechanical ventilation, 3 died and 3 survived (TPE group: 2 recovered and 1 died, Non-TPE group: 1 recovered and 2 died).

An emergency use of TPE in three patients of Covid-19 with ARDS by Zhang L et al^**19**^ also suggested that TPE had an immediate effect on the treatment of the cytokine storm and improvement of PF ratio. Our study also endorses the same fact as most of our patients showed remarkable reduction in Oxygen demand after 1-2 sessions of TPE.

Median duration of hospitalization in patients treated with TPE was 10 day in our study as compared to symptom onset to recovery time of 18 to 25 days in Zhang et al reported cases. This disparity might be because of the fact that our discharge criteria did not include PCR negativity.

Even in PS-matched control group, our study showed a mortality of 38.5% which is significantly lesser as compared with a large retrospective study carried out by Yang et al^**3**^ showing a mortality of 60% in critical disease. The possible reasons might be the inclusion of moderate, severe and critical categories in our PSM cohort rather than only critical cases and the addition of steroids to all treatment groups having evidence of CRS.

Our mortality was higher than shown by Grasselli et al (28%)^**22**^. However, in their study 58% critical patients were still in ICU and not followed up. Nevertheless, epidemiological characteristics such as median age, higher proportion of male population and most common co morbidity (HTN) closely resemble with Grasselli et al study done in Italy over 1591 patients. It has been seen that corona viruses such as severe acute respiratory syndrome (SARS)-CoV and the Middle East respiratory syndrome–CoV (MERS) predominantly affect male gender^**23**^ and may be for same genetic reasons SARS-CoV-2 is also predominantly affecting male population.

Nonetheless, our study had few limitations. Firstly, it was retrospective study which in itself has weaker evidence as compared to prospective trials. Secondly, although a strict PSM analysis was done, but still all biases cannot be neutralized. Thirdly, we did not follow up patients after discharge, due to lack of resources. Fourthly, in TPE procedure, we used centrifugation TPE machine rather than continuous hemofiltration (CHF) which is known to remove IL-6 and similar cytokines molecular mass of 21 to 54.1 kDa^**24**^, due to lack of availability of filters.

Nevertheless, even after considering such limitations, using TPE early, in addition to standard treatment in patients with COVID-19 can mitigate the cytokine storm. TPE shows promise, and we propose that large, multi-centric, randomized trials be designed to further investigate its role.

## CONCLUSION

In conclusion, TPE might be a lifesaving modality, with a statistically significant survival benefit, a decreased hospitalization time and an almost halved CRS resolution time, if started earlier at the onset of CRS in treatment of severe and critical COVID-19.

## Data Availability

All data is available and can be shared on reasonable demand via email

